# Temperature dominates dengue transmission in Thailand: Machine learning reveals critical thresholds and COVID-19 disruption

**DOI:** 10.1101/2025.09.01.25334875

**Authors:** Pikkanet Suttirat, Sudarat Chadsuthi, Supassorn Aekthong, Joacim Rocklöv, Dominique J. Bicout, Peter Haddawy, Saranath Lawpoolsri, Charin Modchang

## Abstract

**Background:** Dengue fever remains a critical public health challenge in Thailand, with transmission dynamics driven by complex interactions between environmental and socioeconomic factors. Understanding these drivers is essential for developing robust prediction systems.

**Methods:** We developed a machine learning framework to classify spatiotemporal dengue risk and identify key drivers of transmission across Thailand. We analyzed 20 years of monthly dengue hemorrhagic fever surveillance data (2003-2022) from 77 provinces, integrating 54 environmental, climatic, and socioeconomic variables. Using eXtreme Gradient Boosting (XGBoost) with SHapley Additive exPlanations (SHAP), we classified provinces as high-risk or low-risk based on the national median incidence. The dataset was stratified into training (2003-2016) and testing periods, with the latter subdivided into pre-COVID-19 (2017-2019), COVID-19 (2020-2021), and post-COVID-19 (2022) phases.

**Results:** The model achieved robust performance with an area under the curve (AUC) of 0.94 during training and 0.80 in pre-pandemic testing. Temperature emerged as the dominant predictor, with temperature-related variables comprising seven of the ten most influential features. Critical transmission thresholds were identified at approximately 21°C for a 1-month lagged minimum temperature and approximately 32°C for a 3-month lagged maximum temperature. Interestingly, precipitation contributed minimally to model predictions, while a higher Gross Provincial Product was associated with an increased risk of dengue, reflecting urban transmission patterns. Model performance deteriorated significantly during the COVID-19 pandemic (AUC = 0.62 in 2021), with systematic overprediction suggesting that behavioral factors outweighed environmental drivers during the pandemic disruption.

**Conclusions:** Temperature, particularly with lags of 1-3 months, is the primary predictor of dengue risk in Thailand. The pandemic-induced disruption of model accuracy underscores the crucial role of human behavioral factors in influencing dengue transmission dynamics. Our results challenge traditional precipitation-focused models and highlight the importance of temperature-driven approaches for dengue prediction in Thailand.

**Plain language summary:** This study used machine learning to predict dengue fever outbreaks across Thailand’s provinces from 2003 to 2022. By analyzing climate data, economic indicators, and satellite imagery, our machine learning model could accurately identify high-risk areas about 80% of the time. We found that temperature is the most important factor determining where and when dengue will spread. Additionally, we identified a critical temperature threshold where dengue transmission essentially stops when minimum temperatures drop below approximately 21°C. Surprisingly, rainfall patterns, which are often emphasized in dengue predictions, played a much smaller role than expected. Higher economic development was also found to be associated with an increased risk of dengue, probably due to urbanization creating ideal conditions for mosquitoes to breed all year round and higher human population density that facilitates virus transmission. However, the COVID-19 pandemic disrupted the model’s accuracy, causing it to predict more dengue cases than actually occurred from 2020 to 2022, likely due to the COVID-19 control measures that climate data alone could not capture. This research demonstrates both the power and limitations of using environmental and socioeconomic data to predict dengue outbreaks in Thailand.

## Introduction

Dengue fever represents one of the most rapidly expanding vector-borne diseases globally, with an estimated 390 million infections occurring annually across more than 100 endemic countries in tropical and subtropical regions ^1–3^. Over the past five decades, the global disease burden has increased dramatically, with a 30-fold rise in incidence ^4^. This escalation is particularly pronounced in Southeast Asia, which bears approximately 70% of the global dengue burden ^1^. Among Southeast Asian nations, Thailand consistently reports some of the highest incidence rates in the region, making it a critical focal point for understanding dengue transmission dynamics ^5^.

Thailand’s experience with dengue dates back to 1958, when the country recorded its first epidemic of dengue hemorrhagic fever (DHF) ^6,7^. Since then, the disease has evolved into a persistent public health challenge, exhibiting complex spatiotemporal patterns driven by the interplay of climatic conditions, socioeconomic factors, and vector ecology ^8,9^. Understanding these patterns is crucial for developing effective prediction and control strategies.

The dengue virus comprises four distinct serotypes and is transmitted to humans primarily through the bites of infected *Aedes* mosquitoes in tropical and subtropical environments ^2,3,10^.

Clinical manifestations range considerably in severity, from asymptomatic infections or mild undifferentiated fever to classical dengue fever (DF), and in severe cases, to life-threatening conditions such as dengue hemorrhagic fever (DHF) or dengue shock syndrome (DSS) ^2,3,11^. The most severe forms can lead to plasma leakage, hemorrhage, and organ impairment, resulting in substantial morbidity and mortality ^11,12^.

The epidemiology of dengue is shaped by a complex interplay of socioeconomic, demographic, and infrastructural factors ^13–15^. While several studies have documented associations between low socioeconomic status and increased local dengue risk ^14,16^, the relationship is more nuanced than initially understood. Rapid urbanization, high population density, and inadequate waste management systems create favorable conditions for disease emergence and spread ^17,18^. However, recent evidence challenges the traditional view of dengue as primarily an urban disease of poverty. Indeed, the virus now affects both rural and urban populations across diverse socioeconomic strata, indicating that transmission patterns have evolved beyond simple associations with urbanization or economic disadvantage ^19,20^.

Environmental factors play a pivotal role in dengue transmission dynamics. Climate-related variables, particularly temperature and rainfall patterns, along with the degree of urban development, fundamentally shape local dengue risk profiles ^1,21,22^. Temperature exerts particularly strong effects on transmission by influencing critical mosquito biological parameters, including biting frequency, fecundity, developmental rates, adult survival, and the efficiency of egg-to-adult development ^23,24^. These temperature-dependent processes create distinct geographic and seasonal patterns of transmission risk.

Traditional approaches to dengue forecasting have relied predominantly on epidemiological surveillance data combined with basic statistical models that incorporate rainfall and temperature variables ^25–27^. However, these conventional methods often fail to capture the complex, non-linear relationships that characterize the interactions between multiple environmental and socioeconomic transmission drivers ^28^. This limitation has prompted researchers to explore more sophisticated analytical approaches.

Recent advances in machine learning offer promising alternatives for disease prediction, with ensemble methods such as gradient boosting demonstrating remarkable performance in capturing intricate feature interactions that traditional models miss ^28–30^. These methods can process high-dimensional data and identify complex patterns that would be difficult to detect using conventional statistical approaches.

Despite growing interest in applying machine learning to dengue prediction, significant knowledge gaps still persist ^29,31^. Most existing models suffer from a limited geographic scope or analyze only short time periods ^29,31,32^. Many rely disproportionately on precipitation as a primary predictor, despite mounting evidence of inconsistent associations between rainfall and dengue incidence ^33^. Additionally, few models provide interpretable insights into which features drive their predictions, limiting their utility for public health decision-making ^28^.

Additionally, the impact of major societal disruptions on the performance of environmentally driven prediction models remains poorly understood, with few studies systematically evaluating how events like the COVID-19 pandemic affect the performance of these models ^34^. Pandemic interventions, including mobility restrictions and lockdown measures, likely altered dengue transmission patterns through mechanisms independent of environmental conditions ^34–37^. The COVID-19 pandemic offers a unique opportunity to assess the model’s resilience and determine the relative contributions of environmental versus behavioral factors in disease transmission. Understanding how prediction systems perform during such extraordinary circumstances is crucial for developing robust forecasting tools that can maintain accuracy in the face of future societal disruptions.

To address these critical gaps, this study develops and evaluates a machine learning framework for spatiotemporal dengue risk classification across Thailand. We employ eXtreme Gradient Boosting (XGBoost) combined with SHapley Additive exPlanations (SHAP) to analyze two decades of dengue surveillance data (2003-2022) integrated with comprehensive environmental and socioeconomic variables. Our specific objectives are threefold: first, to develop a robust classification model capable of quantifying the relative importance and interaction effects of diverse environmental and socioeconomic drivers; second, to identify critical thresholds in key predictors that govern transitions in dengue transmission risk; and third, to evaluate model performance across different epidemiological periods, with particular emphasis on assessing how the COVID-19 pandemic affected prediction accuracy. Through this comprehensive approach, we aim to advance both the theoretical understanding of dengue transmission dynamics and the practical application of machine learning for disease forecasting in complex real-world settings.

## Materials and methods

### Study design and data sources

#### Dengue case data

Data used in this study are monthly dengue hemorrhagic fever (DHF) case reports obtained from the Bureau of Epidemiology, Department of Disease Control, Ministry of Public Health, Thailand, spanning January 2003 to December 2022. Case definitions followed national surveillance standards, where positive cases were identified through either clinical assessment at the point of care or laboratory confirmation at reporting hospitals. Provincial-level data were aggregated monthly, with cases from Buengkarn province incorporated into Nong Khai province after it was administratively separated from Nong Khai in 2011 for consistency. The epidemiological unit is the province, and the time step is the month.

#### Climate variables

Climate data were extracted from the TerraClimate dataset ^38^, which provides high-resolution (4 km^2^) gridded climate information. We obtained six primary variables: precipitation, minimum temperature, maximum temperature, soil moisture, vapor pressure, and wind speed. These gridded data were spatially averaged to the provincial level to match our epidemiological units. Additionally, we derived 19 bioclimatic indicators from monthly temperature and precipitation data using the R package ‘dismo’ ^39,40^. These bioclimatic variables capture annual trends, seasonality, and environmental variability patterns relevant to disease transmission.

#### Socioeconomic indicators

We incorporated three socioeconomic factors to account for urbanization and economic development effects on dengue transmission. Gross Provincial Product (GPP), adjusted for inflation using the Laspeyres Index, was obtained from the National Economic and Social Development Council (https://www.nesdc.go.th). Demographic data, including provincial population statistics and mean household size, were acquired from the Bureau of Registration Administration (https://stat.bora.dopa.go.th). Mean provincial household income data were obtained from the National Statistical Office of Thailand (https://www.nso.go.th), where values are reported biennially with missing years interpolated linearly.

#### Remote sensing data

Environmental conditions were assessed using Moderate Resolution Imaging Spectroradiometer (MODIS) data from the Terra satellite (MOD09A1: Surface Reflectance 8-Day L3 Global 500m SIN Grid V005). We calculated two ecological indices (MNDWI and NDVI) following our established methodology ^41^. Briefly, the water index was derived using the Modified Normalized Difference Water Index (MNDWI) calculated from green and infrared band 7, expressed as the percentage of provincial pixels with MNDWI > 0. The vegetation index was calculated using the Normalized Difference Vegetation Index (NDVI) from red and near-infrared bands, expressed as the percentage of provincial pixels with NDVI values between 0.5 and 1.0. The full description and source of the variables are summarized in **Table 1**.

**Table 1.**
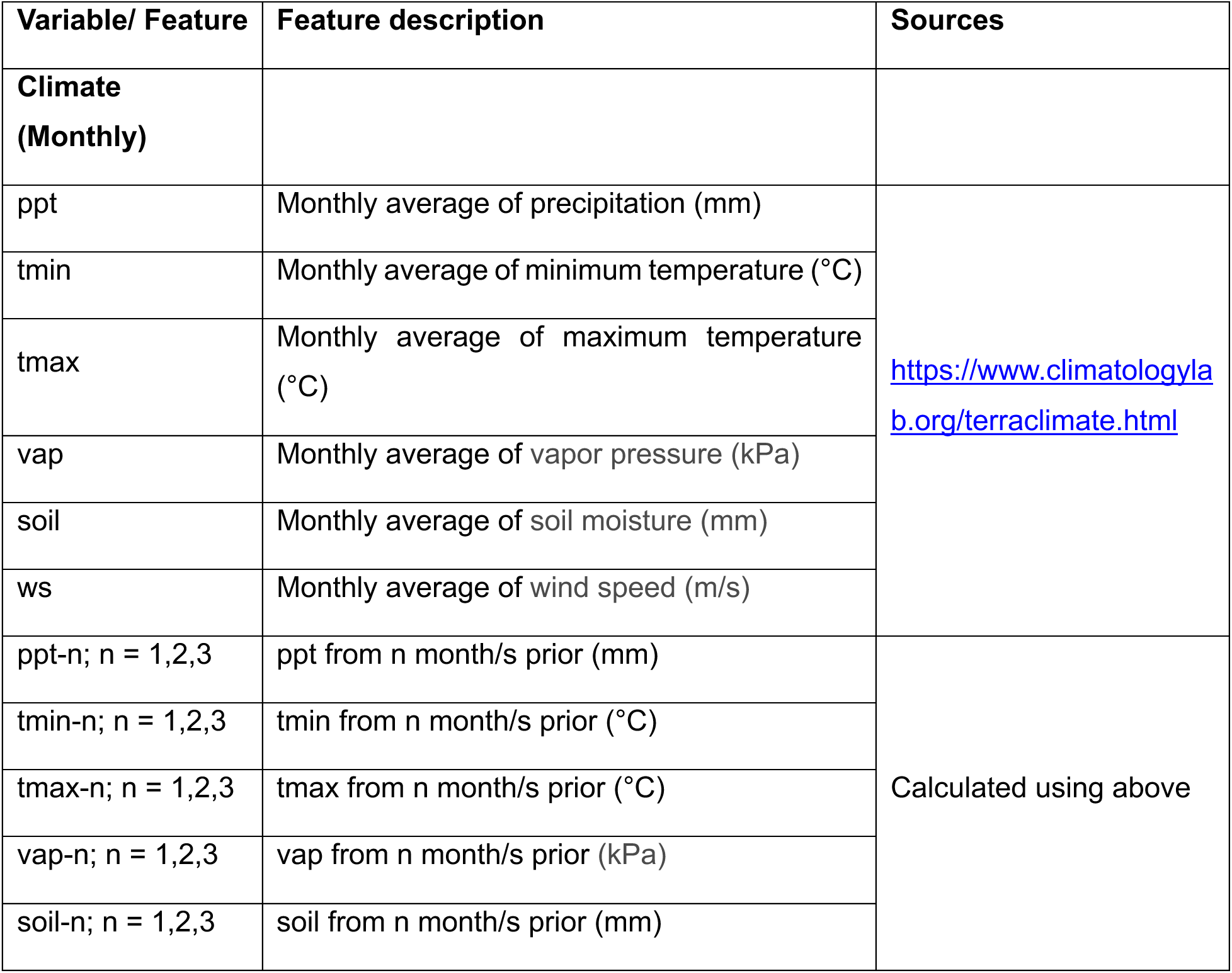
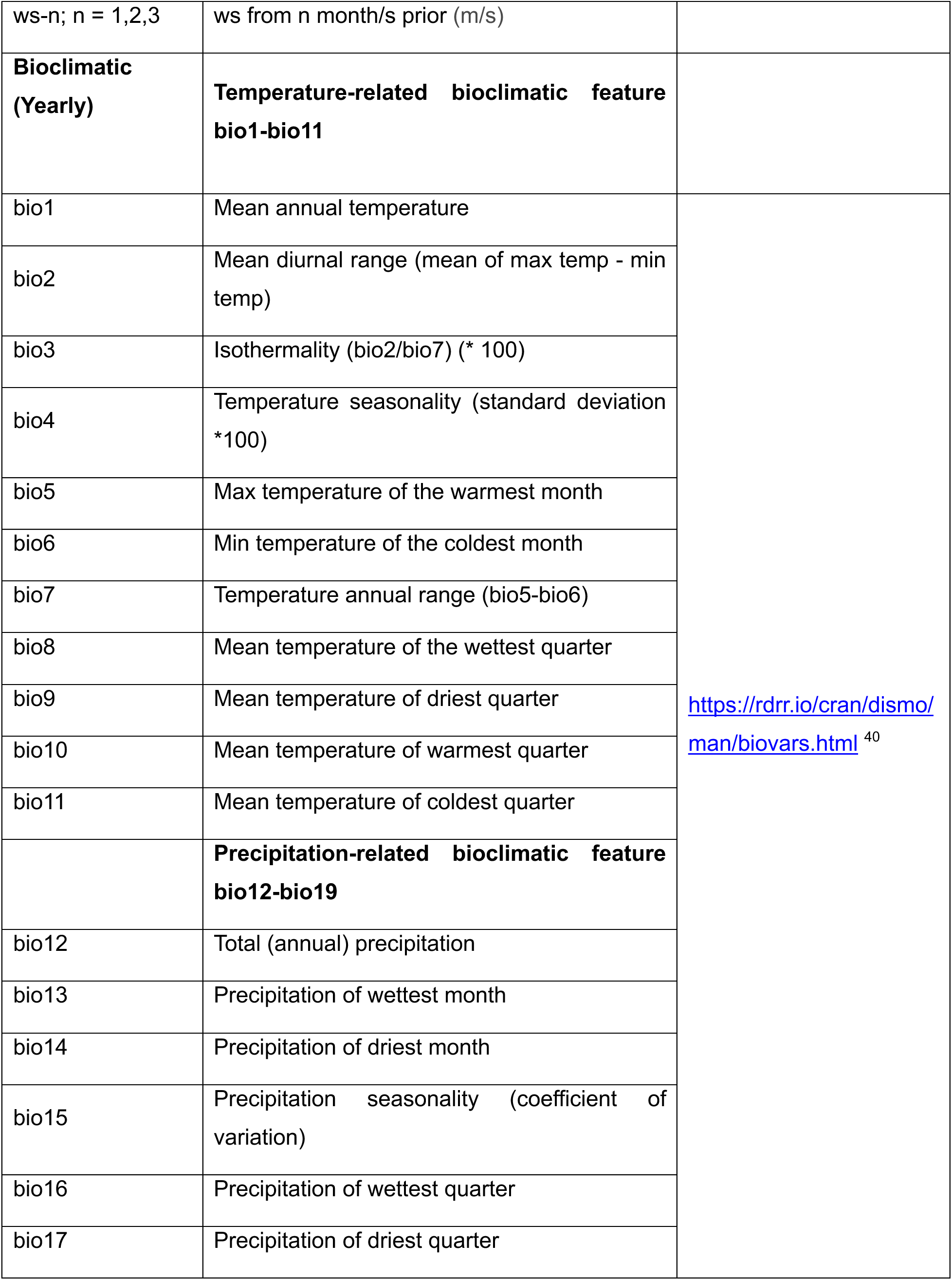
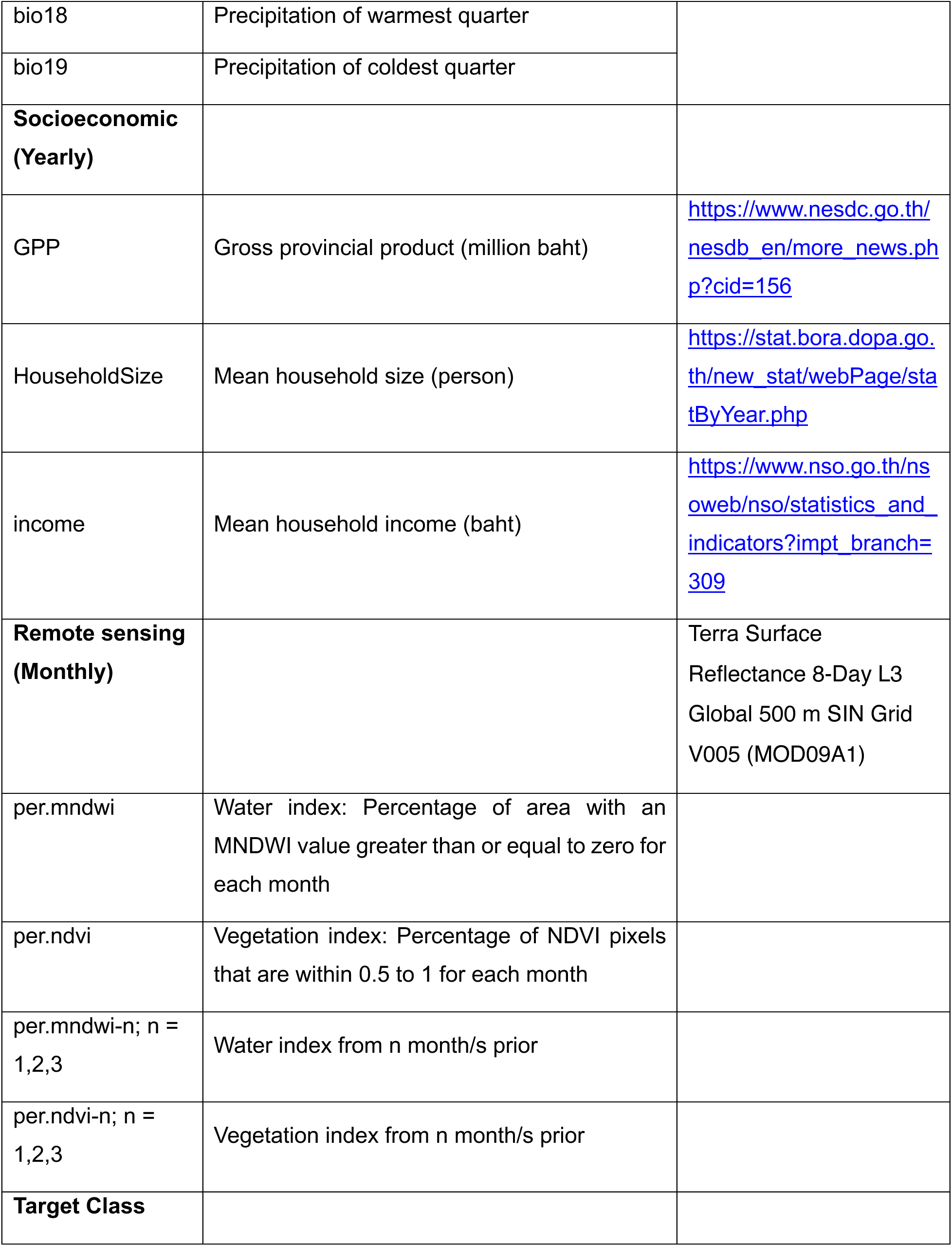
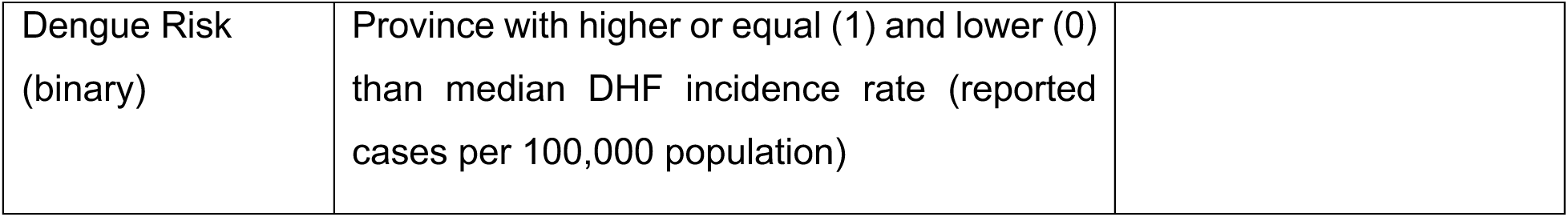
Variables used in the XGBoost model for dengue risk classification in Thailand.

### Data processing and feature engineering

#### Temporal partitioning

We divided the dataset into training (2003-2016, 14 years) and testing (2017-2022, 6 years) periods. This split provided sufficient training data to capture inter-annual variability, including multiple dengue epidemic cycles, while reserving recent years for independent validation. Within the training set, we used the final two years (2015-2016) as a validation set for early stopping to prevent overfitting during model optimization. The testing period was further stratified into three distinct epidemiological phases: pre-COVID-19 (2017-2019), representing baseline model performance under typical transmission conditions; COVID-19 (2020-2021), capturing the period of active pandemic restrictions and interventions; and post-COVID-19 (2022), assessing whether model performance recovered following the relaxation of pandemic measures.

#### Risk classification

Our objective was to develop an operationally relevant early warning system that flags province-months as high versus low dengue risk, rather than predicting exact case counts. Binary classification offers several methodological advantages in this epidemiological context, including providing actionable decision boundaries for public health surveillance and resource allocation. Additionally, binary classification is more robust to systematic reporting biases and temporal non-stationarity than count-based regression models.

We calculated monthly DHF incidence rates per 100,000 population for each province. Using the national median incidence from the training period (2.78 per 100,000) as our classification threshold, we labeled each province-month observation as high-risk (≥ median) or low-risk (< median). This median-based approach ensures balanced target classes (approximately 50% in each category), thereby optimizing machine learning performance and preventing bias toward the majority class. We employed stratified 5-fold cross-validation to maintain this class balance across all training folds, further stabilizing the learning process.

#### Lagged variables

To capture delayed environmental effects on dengue transmission, we generated 1-, 2-, and 3-month lagged values for all climate and remote sensing variables, expanding our feature set to account for temporal dependencies in disease dynamics.

### Machine learning approach

#### XGBoost model implementation

In this work, the supervised machine learning model was employed to capture intricate feature interactions in the training set data and classify each province based on the input features. XGBoost algorithms were selected to solve this classification task. This algorithm constructs an ensemble of decision trees using the combination of gradient boosting tree algorithms and an approximate tree learning technique to distinguish between target classes ^42^. The XGBoost algorithms work based on additive training, where a new tree is constructed to correct the errors made by the previous trees. The new tree was iteratively created to minimize the errors until the errors could not be reduced further, or a specific stop condition was met.

#### Model interpretability

To address the inherent complexity of XGBoost feature interactions, we implemented SHapley Additive exPlanations (SHAP) ^43^. SHAP quantifies each feature’s contribution to individual predictions using cooperative game theory principles. This approach decomposes model predictions into additive feature contributions, enabling both global feature importance ranking and local prediction explanations.

#### Hyperparameter optimization

We optimized model hyperparameters using Bayesian optimization with the Tree-Structured Parzen Estimator (TPE) algorithm implemented in the Python package ‘Optuna’ ^44^. TPE employs performance-aware sampling, concentrating the search in promising hyperparameter regions. The optimization process targeted the number of trees, learning rate, maximum tree depth, subsampling ratio, column sampling by tree, minimum child weight, and regularization parameters (gamma, alpha, lambda). We used 5-fold stratified cross-validation on the training set, preserving class balance in each fold. The optimization objective was to minimize log-loss, with the optimal hyperparameter set selected based on average cross-validation performance. We additionally introduced the early-stopping mechanism to further prevent overfitting the boosted tree to the training set. In this work, we set this early stopping round to 30, using the last two years of the training set as the validation set for the early stopping process.

#### Performance metric

To measure XGBoost models’ performance, an evaluation metric for classification was used. The Area Under the receiver operating characteristic Curve (AUC) was selected to measure the model performance. AUC is designed to measure the total predictive power of the model regardless of the classification threshold. By adjusting the classification threshold, one can observe the tradeoff between two key metrics: true positive rate (sensitivity) and false positive rate (1-specificity). The threshold-dependent metrics were also utilized to additionally measure model performance on XGBoost prediction. The prediction probabilities of XGBoost are transformed to high-risk (class 1) and low-risk (class 0) predictions based on whether the probabilities are higher or lower than the selected threshold, respectively. Four metrics were chosen as follows:

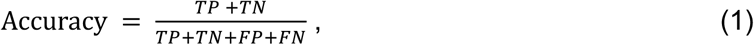

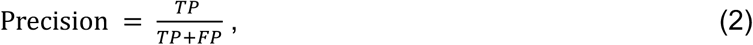

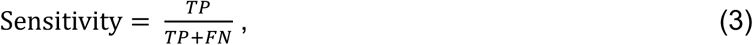

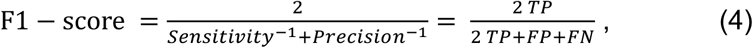

where *TP* is the number of true positive predictions, *TN* is the number of true negative predictions, *FP* is the number of false positive predictions, and *FN* is the number of false negative predictions.

#### Implementation details

All analyses were conducted using Python 3.10 with the following key packages: pandas for data manipulation, numpy for numerical operations, scikit-learn for preprocessing and evaluation, XGBoost for model training, and SHAP for interpretability analysis.

## Results

### Spatiotemporal patterns of dengue hemorrhagic fever in Thailand

Throughout the 20-year study period (2003-2022), Thailand documented 681,430 dengue hemorrhagic fever (DHF) cases, exhibiting substantial spatiotemporal heterogeneity (**Fig. 1a**). Monthly case counts demonstrated dramatic variation, ranging from a maximum of 13,601 cases in July 2013 to a minimum of 105 cases in February 2022. The seasonal distribution revealed pronounced periodicity, with peak transmission occurring during the early wet season, specifically in June (13.16%), July (14.96%), and August (13.98%), totaling 42.12%. In contrast, February consistently showed the lowest disease burden (3.71%) (**Fig. S1**). Among Thailand’s administrative regions, the southern provinces experienced the highest regional incidence rate, reaching 13.93 per 100,000 population in 2010. Notably, the years 2014 and 2016 exhibited markedly reduced DHF incidence compared to other years within the training period, suggesting the potential influence of climatic anomalies or intensified control measures.

**Figure 1.**
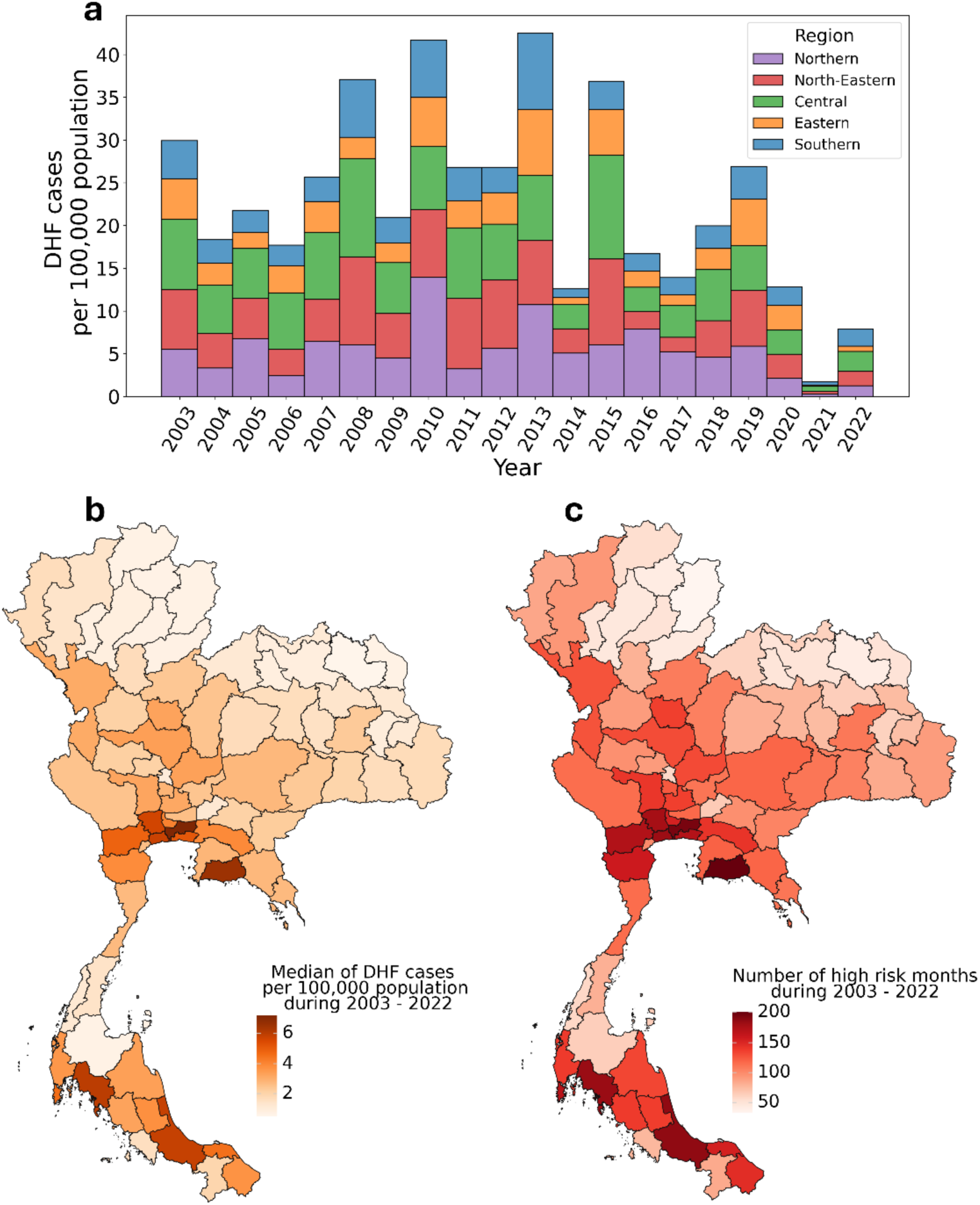
Spatiotemporal distribution of dengue hemorrhagic fever (DHF) in Thailand, 2003–2022. **(a)** Annual DHF incidence rates by region, showing temporal trends across Thailand’s five major administrative regions. **(b)** Geographic distribution of median provincial DHF incidence rates over the study period, with color intensity corresponding to disease burden (darker shades indicate higher incidence per 100,000 population). **(c)** Cumulative number of high-risk months by province during 2003 – 2022, where high risk is defined as monthly incidence exceeding the national median threshold of 2.78 per 100,000 population (derived from 2003–2016 training data). Provinces with higher values indicate persistent dengue hotspots.

Geographic analysis identified persistent high-risk zones predominantly concentrated in central and southern Thailand **(Fig. 1b**). Utilizing the national median incidence rate of 2.78 cases per 100,000 population as a binary classification threshold, we delineated provinces with sustained elevated dengue burden. The cumulative frequency analysis of high-risk months revealed that certain provinces maintained endemic transmission for extended periods (**Fig. 1c**). This epidemiological threshold subsequently served to categorize each province-month observation as either high-risk or low-risk, establishing the target classes for our machine learning classification framework.

### Model performance and validation

The XGBoost model demonstrated robust discriminative capacity, achieving an area under the receiver operating characteristic curve (AUC) of 0.94 on the training dataset (2003-2016). Model performance remained consistently strong during the pre-COVID-19 testing period (2017-2019), maintaining an AUC of 0.80. However, performance deteriorated markedly during and following the COVID-19 pandemic (2020-2022), declining to an AUC of 0.74 (**Fig. 2a**). Complementary threshold-dependent metrics, including accuracy, precision, sensitivity, and F1-score, corroborated the model’s ability to effectively discriminate between high and low-risk provinces throughout the pre-pandemic period (2003-2019) (**Fig. S2**).

**Figure 2.**
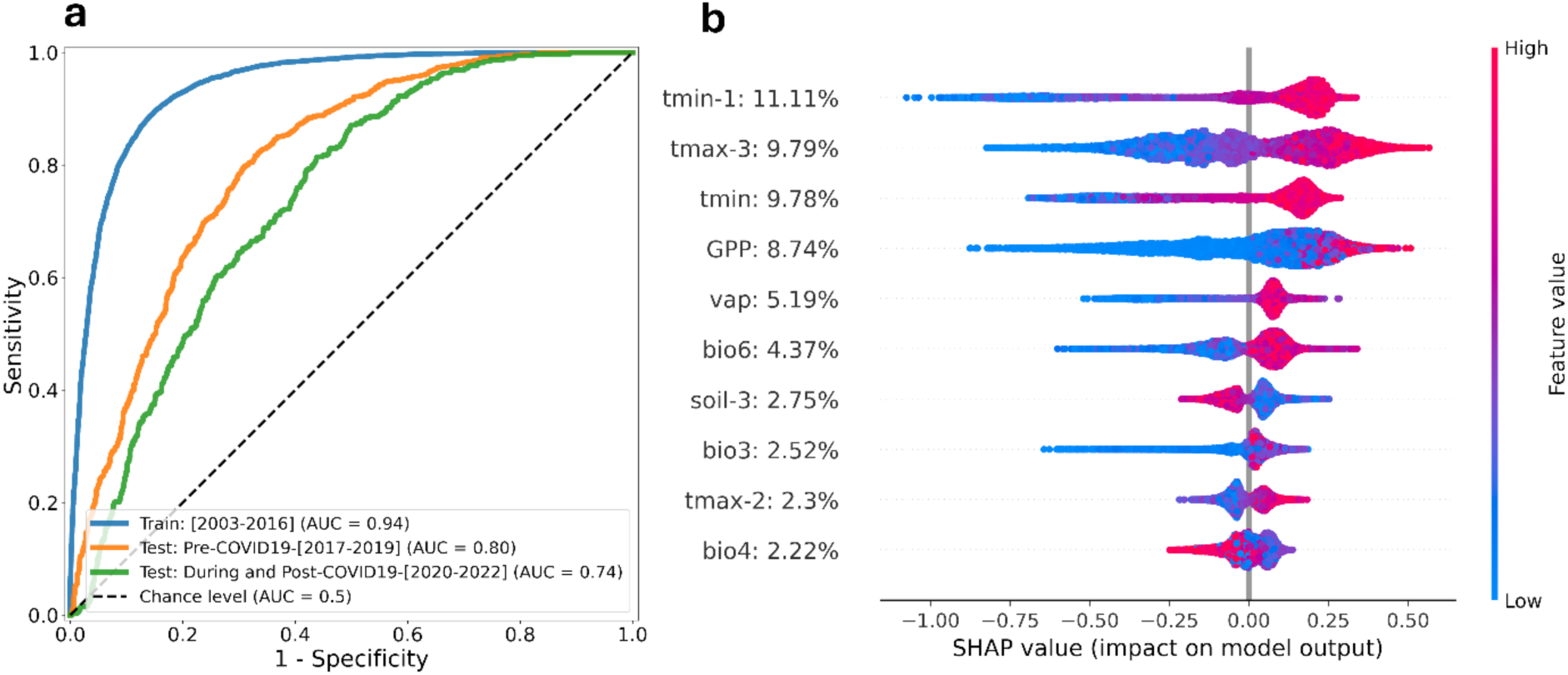
XGBoost model performance and feature importance analysis. **(a)** Receiver Operating Characteristic (ROC) curves comparing model discrimination across training (2003– 2016) and test datasets (2017–2022), with Area Under the Curve (AUC) values indicating predictive performance. The diagonal reference line represents random classification (AUC = 0.5). Model hyperparameters were optimized using 5-fold cross-validation. **(b)** SHAP feature importance analysis showing the top 10 predictive variables ranked by mean absolute SHAP values, based on 2003–2019 data. Percentages indicate each feature’s relative contribution among all 54 input features. Each dot represents an individual observation, colored by feature value (red = high, blue = low). Horizontal position indicates the feature’s impact on predicted dengue risk, with positive SHAP values increasing and negative values decreasing the probability of high-risk classification.

Temporal stratification of model predictions revealed nuanced performance patterns. While the model could accurately capture dengue risk dynamics prior to 2017, a systematic tendency toward overprediction emerged during 2017-2019 (**Fig. 3**). This overprediction pattern suggests the presence of factors influencing dengue transmission that were not captured by our environmental and socioeconomic predictors, such as human mobility patterns ^45–48^. The overprediction bias intensified dramatically following 2020, coinciding with the onset of the COVID-19 pandemic restrictions ^37,45,49^. However, despite these temporal variations in performance, the model maintained an overall accuracy of 79.39% across the entire 20-year study period.

**Figure 3.**
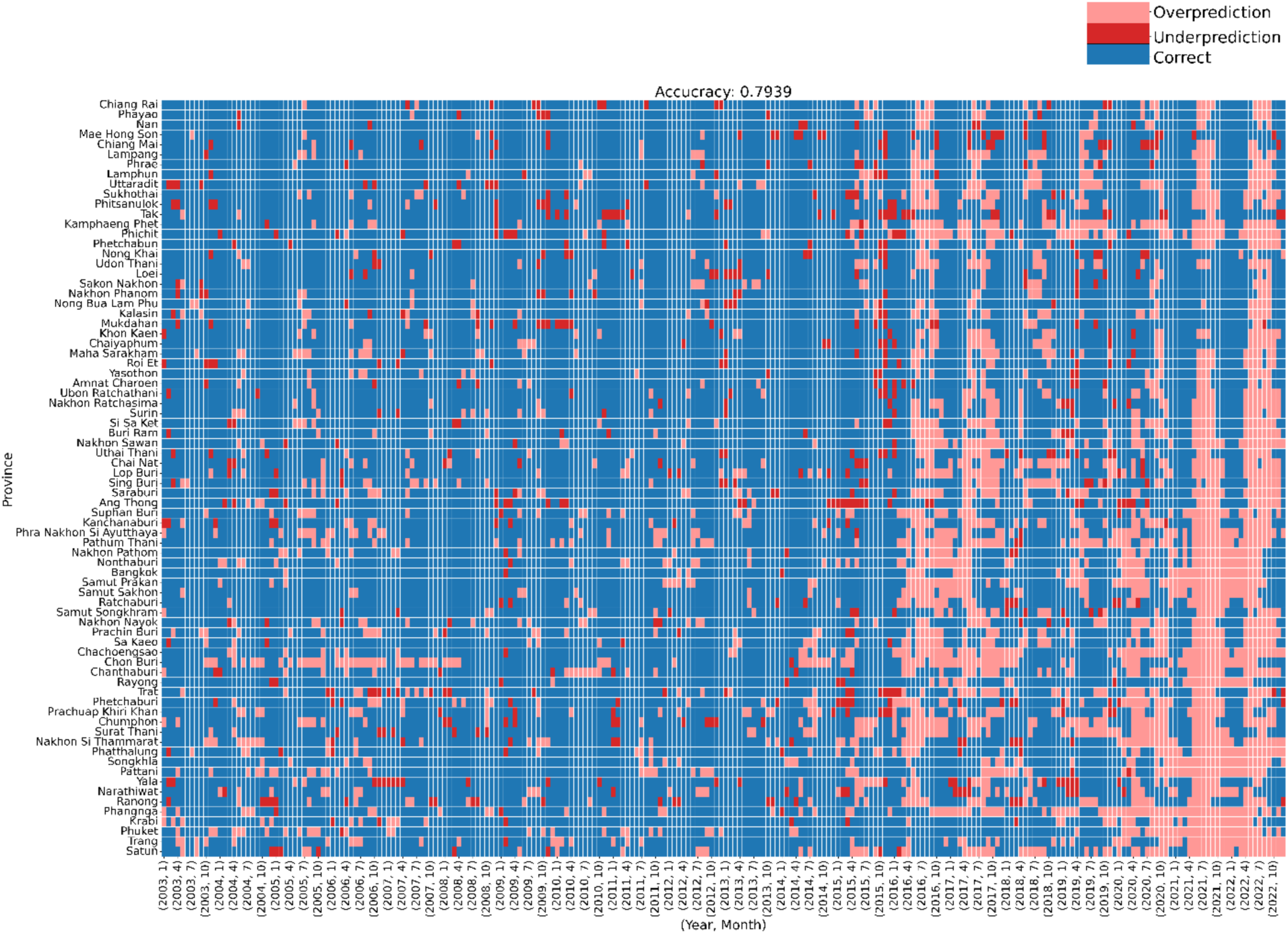
Spatiotemporal evaluation of XGBoost model predictions across Thai provinces, 2003–2022. The heatmap displays monthly prediction accuracy for each province throughout training (2003–2016) and testing (2017–2022) periods. Colors indicate prediction outcomes: correct predictions (blue), false positives/overprediction (pink; high risk predicted when actual risk was low), and false negatives/underprediction (red; low risk predicted when actual risk was high); the ground truth is shown in **Fig. S3**. Provinces are grouped by region and ordered from north to south by latitude. The overall model accuracy is 79.39%.

### Feature importance and interaction analysis

SHAP analysis revealed temperature variables as the dominant predictive features, with remarkable consistency across multiple metrics. Among the 54 input features, temperature-related variables occupied seven of the ten highest-ranked positions based on mean absolute SHAP values (**Fig. 2b**). Despite high correlations among temperature-related features (**Fig. S4**), XGBoost’s tree-based algorithm effectively handled this multicollinearity by selecting the most informative predictors at each split. The two most influential predictors were 1-month lagged minimum temperature (tmin-1) and 3-month lagged maximum temperature (tmax-3), underscoring the critical importance of antecedent thermal conditions in dengue transmission dynamics. The current month’s minimum temperature ranked third, highlighting the combined influence of both immediate and lagged temperature effects on disease risk.

All temperature variables exhibited positive associations with dengue risk across all temporal scales, reinforcing the established biological relationship between elevated temperatures and enhanced dengue transmission potential. Gross provincial product (GPP) emerged as the fourth most influential predictor, with higher economic development associated with increased dengue risk, which reflects the predominantly urban nature of dengue transmission in Thailand. The top ten features collectively accounted for 58.76% of the model’s total predictive capacity. Among these leading predictors, three bioclimatic indices—namely, minimum temperature of the coldest month (bio6), isothermality (bio3), and temperature seasonality (bio4)—were all temperature-derived metrics. In marked contrast, precipitation variables, precipitation-related bioclimatic indices, and remote sensing indicators contributed minimally to model predictions.

SHAP dependence plots illuminated complex nonlinear relationships and significant feature interactions governing dengue risk (**Fig. 4**). The relationship between 1-month lagged minimum temperature (tmin-1) and dengue risk exhibited a distinct threshold effect: temperatures below approximately 21°C were negatively associated with transmission risk, while the positive effect reached a plateau at approximately 24°C. This relationship was substantially modulated by 3-month lagged maximum temperature, with elevated tmax-3 values amplifying the effect of tmin-1 on predicted risk (**Fig. 4a**). Similarly, tmax-3 demonstrated a nonlinear response with a critical threshold at approximately 32°C, beyond which dengue risk increased markedly. This temperature effect was modified by concurrent minimum temperature, with lower tmin values likely attenuating the positive influence of tmax-3, particularly within the 28-31°C range (**Fig. 4b**).

**Figure 4.**
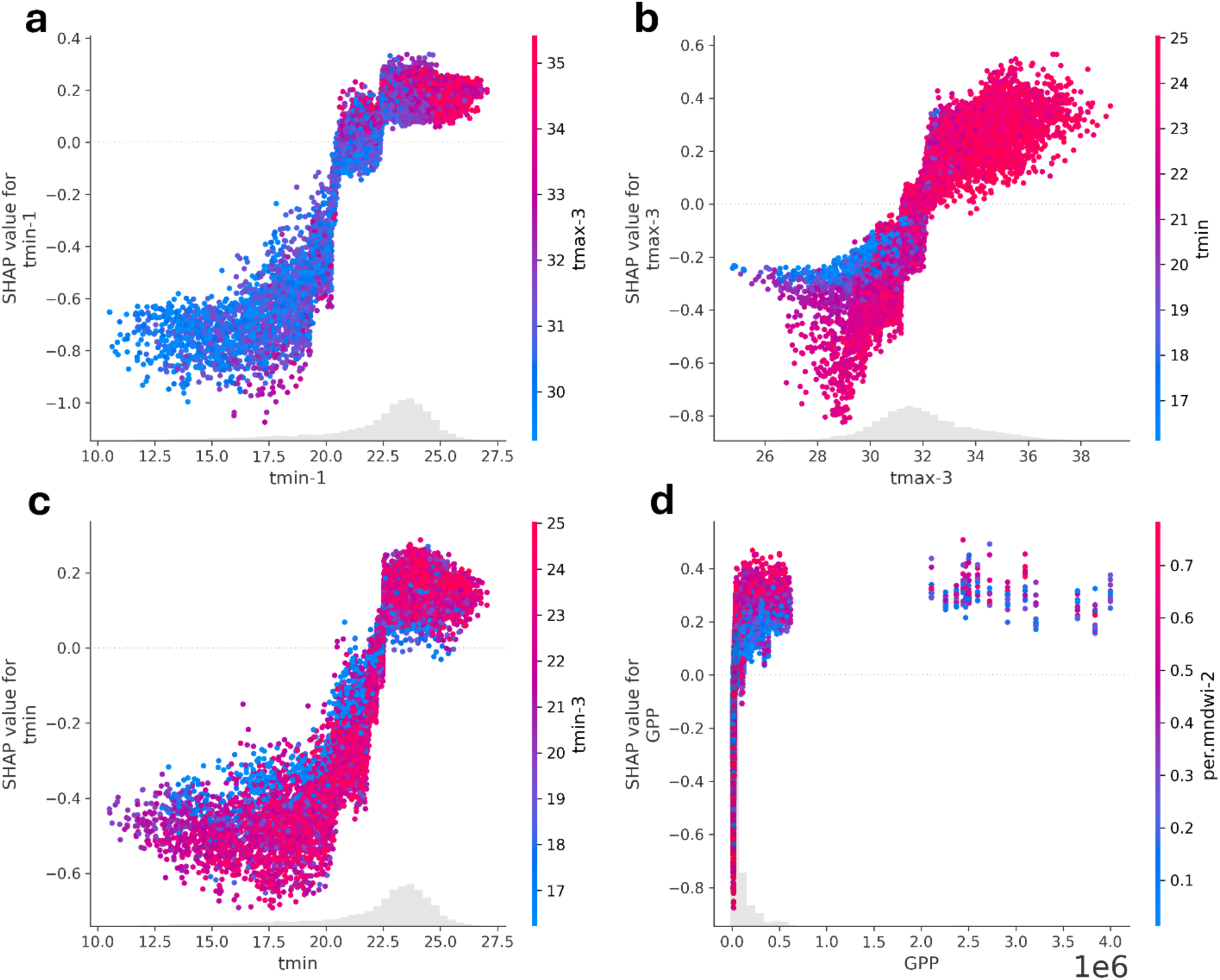
SHAP dependence plots for the four most important predictive features. Each dot represents a single observation (province-month), with the x-axis showing the feature value and the y-axis showing its SHAP value (contribution to predicted log odds). Gray shading indicates the feature distribution across the dataset (2003–2019). Dot color represents the value of the most strongly interacting feature, as determined by SHAP’s interaction analysis, revealing how feature combinations influence dengue risk predictions. Positive SHAP values indicate increased predicted risk, while negative values indicate decreased risk.

The interaction between current and 3-month lagged minimum temperatures revealed a biologically significant threshold at approximately 22°C, potentially representing a fundamental constraint for sustained dengue virus transmission (**Fig. 4c**). The 2-month lagged water index (per.mndwi-2) likely exhibited synergistic interactions with GPP, particularly pronounced in provinces where GPP values remained below 10^6^ million baht. Conversely, this amplifying effect diminished substantially in Bangkok, where GPP values (> 2 x 10^6^ million baht) exceeded those of all other provinces (**Fig. 4d**).

### Impact of the COVID-19 pandemic on dengue dynamics

The COVID-19 pandemic fundamentally disrupted dengue transmission patterns ^49^, resulting in precipitous declines in model performance. Annual AUC reached its lowest at 0.62 in 2021, representing the poorest discriminative capacity observed throughout the entire study period (**Fig. 5a**). This performance degradation persisted into the post-pandemic period, suggesting enduring alterations to dengue epidemiological dynamics. All performance metrics exhibited consistent deterioration during this period, confirming the comprehensive nature of the disruption **(Fig. 5b**).

**Figure 5.**
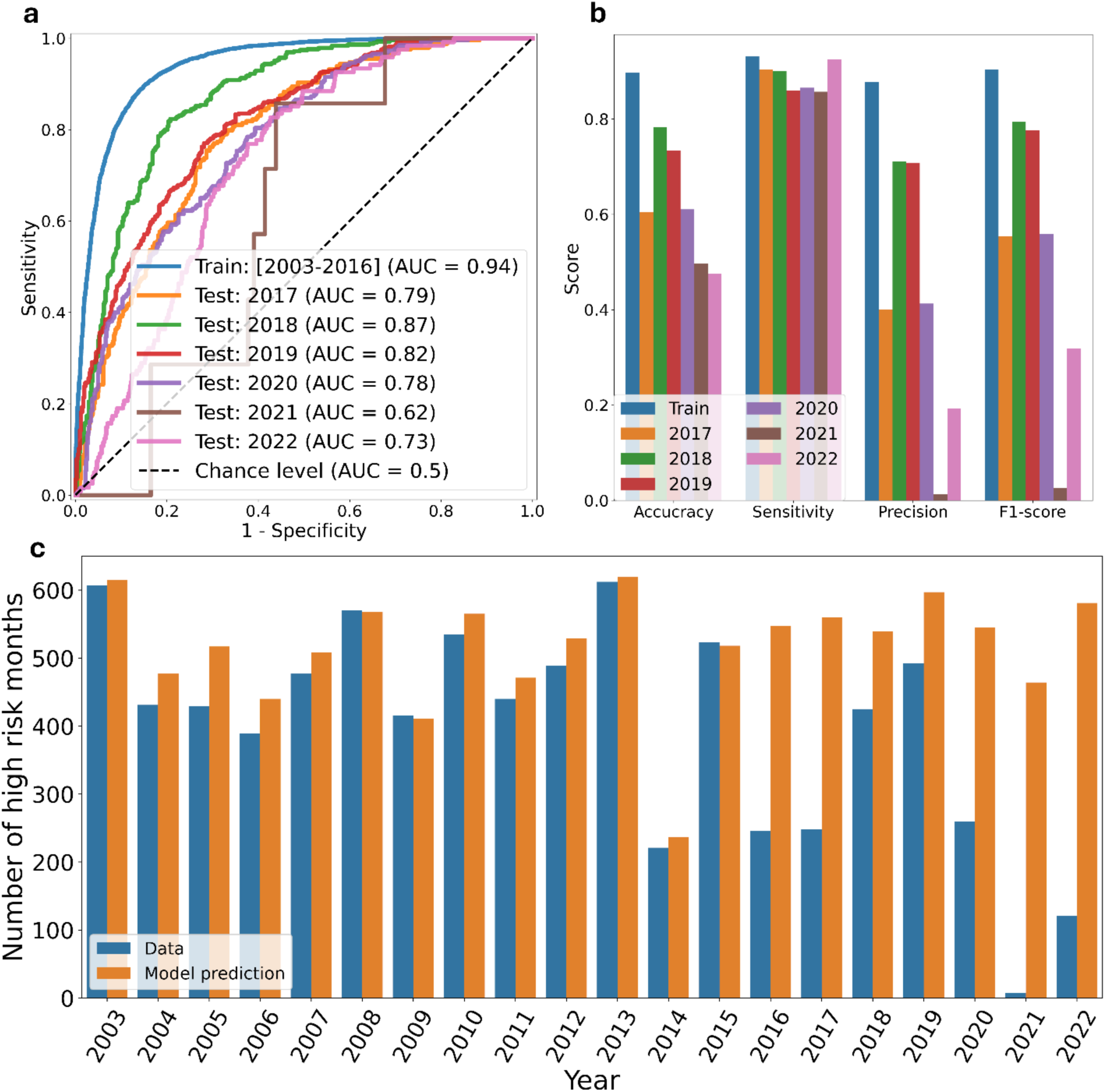
Impact of COVID-19 on model performance across three periods: pre-pandemic (2003–2019), pandemic (2020–2021), and post-pandemic (2022). **(a)** Receiver Operating Characteristic (ROC) curves showing declining model discrimination ability across periods, with AUC values dropping from pre-pandemic baseline. The diagonal line indicates random classification (AUC = 0.5). **(b)** Annual performance metrics (accuracy, precision, sensitivity, and F1-score) calculated using a 0.5 classification threshold, revealing substantial performance degradation during and after the pandemic. **(c)** Comparison of predicted versus observed high-risk province-months by period, demonstrating systematic model overprediction during the pandemic when dengue transmission was suppressed.

The model’s overprediction during the pandemic was substantial. Employing a default classification threshold of 0.5, the model predicted 545, 464, and 581 high-risk province-months for 2020, 2021, and 2022, respectively. In contrast, observed data revealed only 260, 7, and 121 actual high-risk instances during these corresponding periods (**Fig. 5c**). This systematic overestimation indicates that pandemic-related factors, including mobility restrictions, behavioral modifications, and altered healthcare-seeking patterns, suppressed dengue transmission through mechanisms operating independently of climatic drivers ^49,50^. The persistence of this suppression effect into 2022 suggests that dengue transmission dynamics might have undergone fundamental restructuring, necessitating comprehensive model recalibration to restore predictive accuracy.

## Discussion

Our investigation demonstrates the potential of machine learning approaches, particularly eXtreme Gradient Boosting (XGBoost), for predicting dengue hemorrhagic fever risk across Thailand by integrating comprehensive climatic, environmental, and socioeconomic variables. The model achieved robust predictive performance, with an AUC of 0.94 during training and 0.80 in pre-pandemic testing periods, while establishing temperature-related variables as the predominant predictors of dengue transmission risk. Notably, the COVID-19 pandemic’s profound disruption of model performance highlights the complex interplay between environmental drivers and other factors not captured in our model, such as human behavioral factors, disease control measures, and healthcare-seeking behavior, that govern dengue epidemiology in Thailand.

### Temperature as the primary driver of dengue risk

Our analysis indicates that temperature is the most critical environmental determinant of dengue risk throughout Thailand, with temperature-related variables comprising seven of the ten most influential predictive features. This finding aligns with well-established biological mechanisms that govern dengue virus transmission dynamics ^51,52^. Temperature exerts multifaceted effects on the transmission cycle, modulating the extrinsic incubation period of the virus within mosquito vectors, accelerating mosquito developmental rates, and increasing biting frequency within specific thermal ranges ^51–53^.

The identification of a critical threshold effect at approximately 21°C for minimum temperature corresponds remarkably well with established biological constraints. Below this temperature, dengue virus replication within *Aedes aegypti* mosquitoes becomes negligible ^51,52,54^. Indeed, a laboratory study demonstrated that dengue transmission ceases below approximately 18°C due to insufficient viral replication within the mosquito vector at these lower temperatures^54^.

The analysis revealed that the 3-month lagged maximum temperature (tmax-3) exhibited a distinct critical threshold near 32°C. Below this threshold, the effect on dengue risk remained minimal; however, risk increased substantially once monthly maximum temperatures exceeded this point. This threshold likely represents the optimal temperature range where both mosquito development rates and virus replication achieve maximum efficiency ^51,52,55^. Interestingly, the plateau effect observed for maximum temperature was less pronounced compared to that of minimum temperature. This asymmetry suggests that while extremely high maximum temperatures may limit transmission through increased mosquito mortality or reduced virus transmission probability, these constraining effects are less significant than those imposed by lower temperature thresholds ^56^. Such asymmetric temperature effects carry important implications for understanding climate change impacts on dengue transmission, as warming trends may expand suitable transmission zones primarily through elevated minimum temperatures rather than through alterations in maximum temperature patterns ^57^.

The temporal dynamics of dengue transmission are effectively captured by the prominence of lagged temperature variables, particularly 1-month lagged minimum temperature (tmin-1) and 3-month lagged maximum temperature (tmax-3). The 1-month lag for minimum temperature likely reflects immediate effects on adult mosquito survival and viral replication efficiency, whereas the 3-month lag for maximum temperature may represent cumulative impacts on mosquito population dynamics and the temporal requirements for epidemic establishment following favorable environmental conditions ^51,58,59^. These observations align with previous research demonstrating that temperature effects on dengue transmission operate across multiple temporal scales, influencing both immediate vector competence and longer-term population dynamics ^51,58–60^.

Our results indicate that precipitation variables played a much smaller role in model predictions than expected, with the 1-month lagged precipitation variable ranking 11^th^ out of 54 considered factors. While this finding contradicts the traditional emphasis on rainfall in dengue prediction models, it aligns with emerging evidence suggesting that precipitation-dengue relationships are highly context-dependent and often non-monotonic and non-linear ^61^. In Thailand’s urban environments, artificial water containers might provide year-round breeding sites that would remain independent of natural rainfall patterns ^62^. In rural areas, the relationship is equally complex: once precipitation reaches a certain threshold, additional rainfall might only slightly contribute to dengue risk. In fact, Thailand experiences higher average rainfall in August and September than in July, yet has fewer dengue cases **(Figs. S1** and **S5)**. This might be due to excessive rainfall that can flush larvae from breeding containers, thereby reducing vector populations. In contrast, drought conditions may lead to increased household water storage practices, paradoxically creating more breeding opportunities within households ^53,63,64^. This complexity underscores the limitations of simplistic rainfall-based prediction approaches and highlights the need for more nuanced environmental indicators in dengue forecasting models in Thailand.

### Socioeconomic factors and urbanization

The emergence of Gross Provincial Product (GPP) as the fourth most influential predictor reveals a seemingly paradoxical relationship wherein higher economic development correlates with increased dengue risk. This relationship illuminates the fundamentally urban nature of dengue transmission within Thailand. The urbanization process might create optimal conditions for *Aedes aegypti* proliferation through multiple mechanisms: the proliferation of artificial water containers that serve as all-year-round breeding sites, increased human population density and mobility that facilitates virus transmission, and urban heat island effects that maintain temperatures within favorable ranges for vector survival and development ^61,65–67^.

The detected interaction between GPP and environmental variables, particularly the water index, indicates that urbanization fundamentally modifies the relationship between environmental conditions and dengue risk. This modification likely occurs through alterations in mosquito breeding site availability and human behavioral patterns that influence vector-human contact rates ^53,61^. These findings underscore the importance of considering socioeconomic development patterns when designing dengue control strategies in rapidly urbanizing regions.

### Impact of the COVID-19 pandemic

The decline in model performance during the COVID-19 pandemic period, with AUC dropping to 0.62 in 2021, suggests the importance role of human behavioral factors in dengue transmission dynamics. The model’s systematic overprediction throughout this period indicates that pandemic-related interventions, including mobility restrictions, altered healthcare-seeking behaviors, and modified work and educational patterns, suppressed dengue transmission through mechanisms independent of climatic conditions ^45–49,68^.

The persistence of reduced model performance extending into 2022 suggests potential long-term alterations in dengue epidemiology. These changes might encompass shifts in population immunity patterns, and modifications in vector population dynamics. This finding carries crucial implications for dengue prediction models, demonstrating that purely environmental models may fail catastrophically during periods of significant societal disruption ^49,68^. Future modeling efforts should therefore consider incorporating real-time mobility data, healthcare utilization patterns, and other behavioral indicators to maintain predictive accuracy during unprecedented circumstances.

### Limitations

Several limitations warrant careful consideration when interpreting our findings. First, our dependence on passive surveillance data likely results in systematic underestimation of true dengue incidence, particularly in regions with limited healthcare access or reduced healthcare-seeking behavior ^69^. The exclusive use of reported DHF cases rather than comprehensive dengue case data may introduce bias toward more severe disease patterns, potentially limiting the generalizability of our predictions to mild or asymptomatic infections. Second, the absence of direct measures of vector abundance, human mobility, and immunity patterns limits our understanding of transmission dynamics. Third, the effects of dengue control measures were not considered in this study, which might interfere with the dynamics of dengue transmission. Fourth, our binary classification may achieve high accuracy partly by identifying provinces with consistent endemic levels rather than temporal variations. However, since our primary objective was identifying environmental and socioeconomic drivers rather than spatiotemporal prediction, this limitation does not undermine our key findings. Finally, the model’s reduced performance during the COVID-19 pandemic highlights its vulnerability to unprecedented societal changes not captured in the training data.

## Conclusions

Our XGBoost model with SHAP interpretability achieved robust performance for dengue risk prediction in Thailand (training AUC = 0.94, pre-pandemic test AUC = 0.80), demonstrating accurate provincial-level classification using environmental and socioeconomic predictors. Temperature emerged as the dominant driver, accounting for seven of the top ten features, with critical thresholds at approximately 21°C for 1-month lagged minimum temperature and approximately 32°C for 3-month lagged maximum temperature. The prominence of 1-month and 3-month temperature lags emphasizes that thermal history, rather than current conditions, drives transmission risk. We found that higher Gross Provincial Product increased dengue risk, and precipitation contributed minimally to model predictive power, confirming the fundamentally urban-centric nature of transmission patterns in Thailand. The COVID-19 pandemic period revealed critical limitations in environmentally driven prediction models. Model performance declined during this period, with AUC falling to 0.62 in 2021, accompanied by systematic overprediction of risk. This deterioration demonstrates how behavioral modifications can comprehensively override environmental drivers of transmission. These findings underscore the need for adaptive modeling approaches that integrate continuous environmental monitoring with real-time behavioral surveillance to maintain predictive accuracy during periods of societal disruption.

## Data Availability

All data produced in the present study are available upon reasonable request to the authors.

## Acknowledgements

This research has received funding support from The Rockefeller Foundation under the Strengthening the Early Warning and Outbreak Detection Systems through Nation-wide Event-based and Syndromic Surveillance (STONES) project (Grant No. 2021PPI005). Pikkanet Suttirat acknowledges support from the Development and Promotion of Science and Technology Talents Project (DPST) of Thailand.

## Ethics statement

This study received an exemption from ethical review by the Ethics Committee of the Faculty of Tropical Medicine, Mahidol University, Thailand (Documentary Proof of Exemption: MUTM-EXMPT 2024-005, dated 8 July 2024). The exemption was granted as this research involved retrospective analysis of anonymized, aggregated surveillance data with no direct human subject involvement. The Ethics Committee certified that the study complies with the Declaration of Helsinki, ICH Guidelines for Good Clinical Practice, and other international guidelines for human research protection.

