## Supplementary Materials for "Temperature dominates dengue transmission in Thailand: Machine learning reveals critical thresholds and COVID-19 disruption"

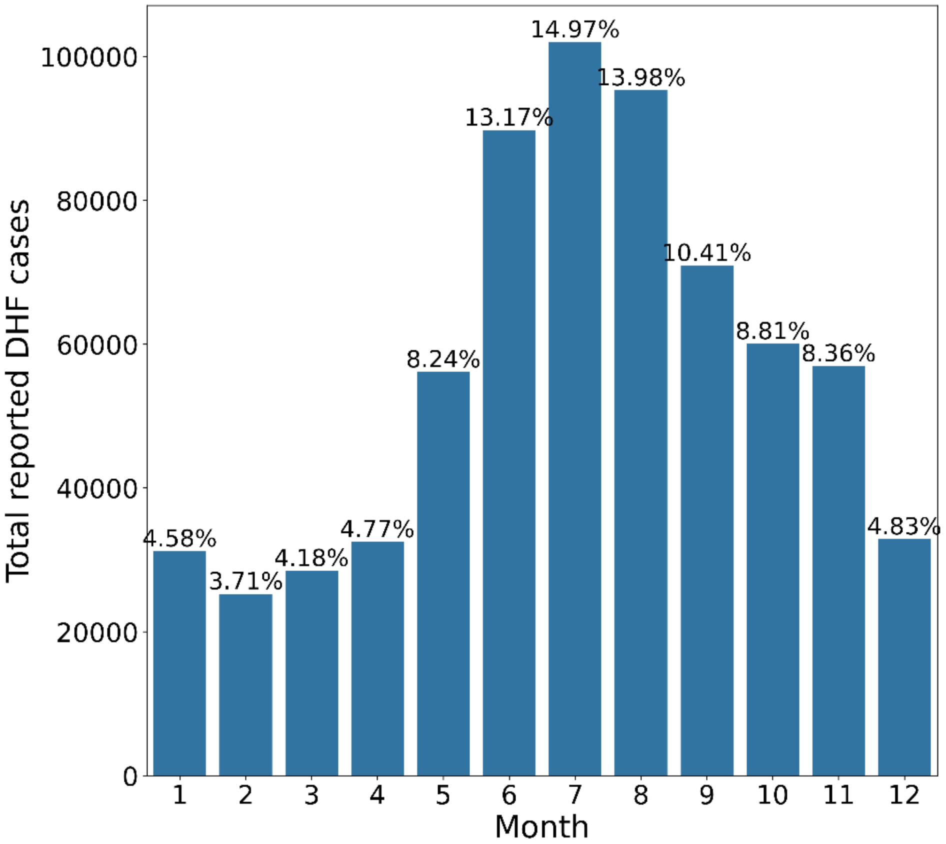


**Figure S1**. Seasonal distribution of dengue hemorrhagic fever cases in Thailand, 2003–2022. Monthly case counts are shown as percentages of total reported cases, revealing peak transmission during the rainy season (June–August) and lowest transmission in the dry season (February).

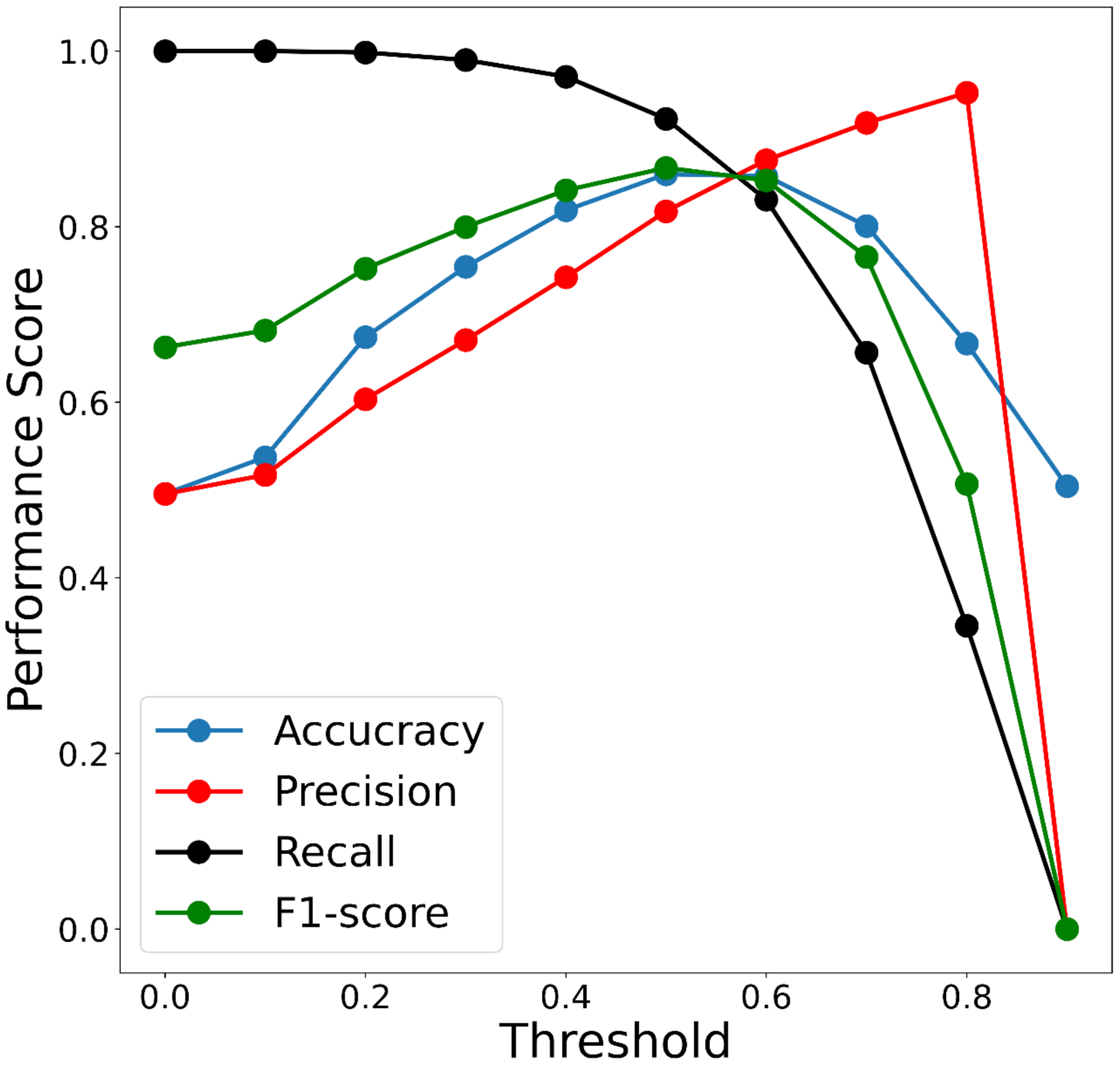


**Figure S2.** Model performance sensitivity to classification threshold, 2003–2019. Four performance metrics (accuracy, precision, sensitivity, and F1-score) are evaluated across probability thresholds from 0 to 1. The threshold determines the predicted probability cutoff for classifying provinces as high-risk versus low-risk.

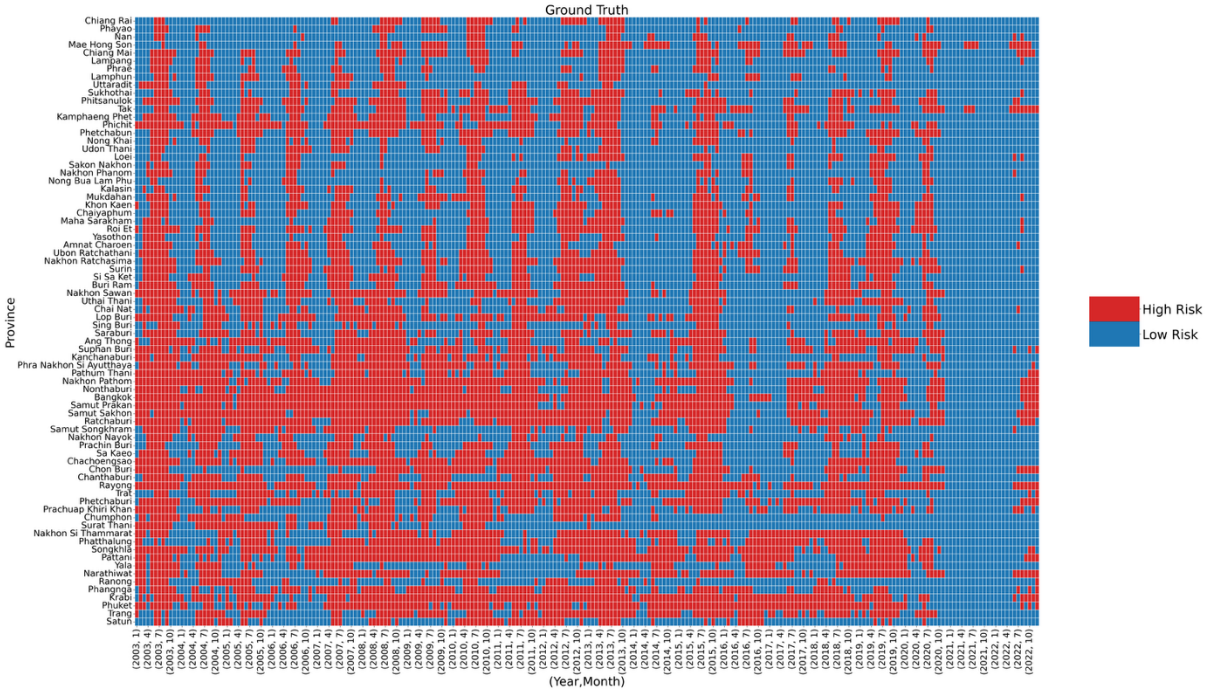


**Figure S3. Ground truth classification of dengue risk across Thai provinces, 2003-2022.** Heatmap showing the actual monthly risk classification for each province based on whether DHF incidence exceeded the national median threshold (2.78 per 100,000 population). Red cells indicate high-risk months (≥ median incidence) and blue cells indicate low-risk months (< median incidence). Provinces are arranged by region and ordered from north to south. This ground truth data serves as the target variable for model training (2003-2016) and validation (2017-2022). Notable patterns include persistent high-risk status in central provinces (notably Bangkok and surrounding areas), while northern provinces show more sporadic high-risk periods. The dramatic reduction in high-risk observations during 2021-2022 reflects the impact of the COVID-19 pandemic on dengue transmission.

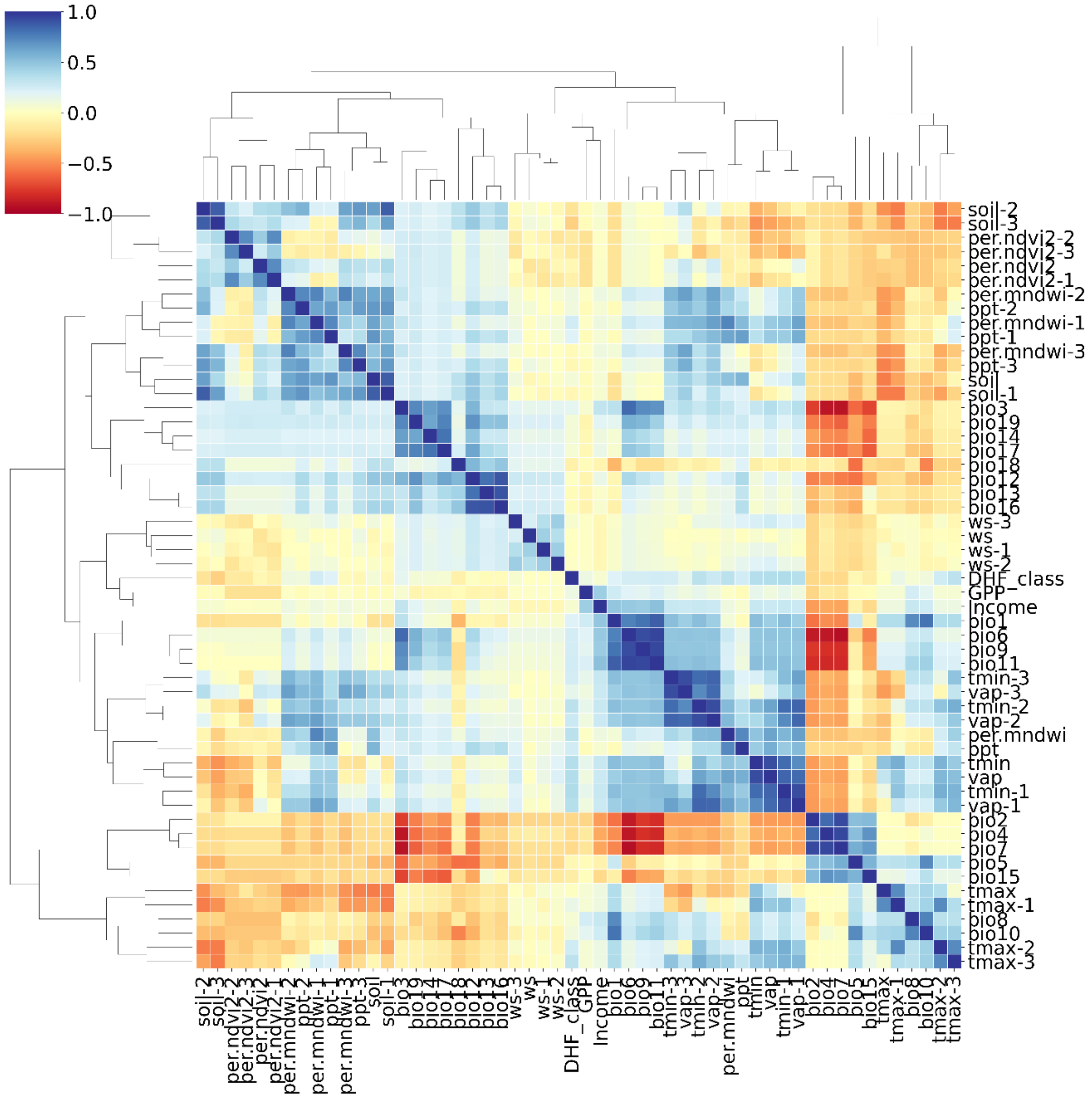


**Figure S4.** Correlation structure and multicollinearity among predictive features. Hierarchically clustered heatmap showing Pearson correlation coefficients between all 54 model features, with color intensity indicating correlation strength (red = negative, blue = positive). Dendrograms reveal natural groupings of related variables, with temperature-related features forming distinct clusters.

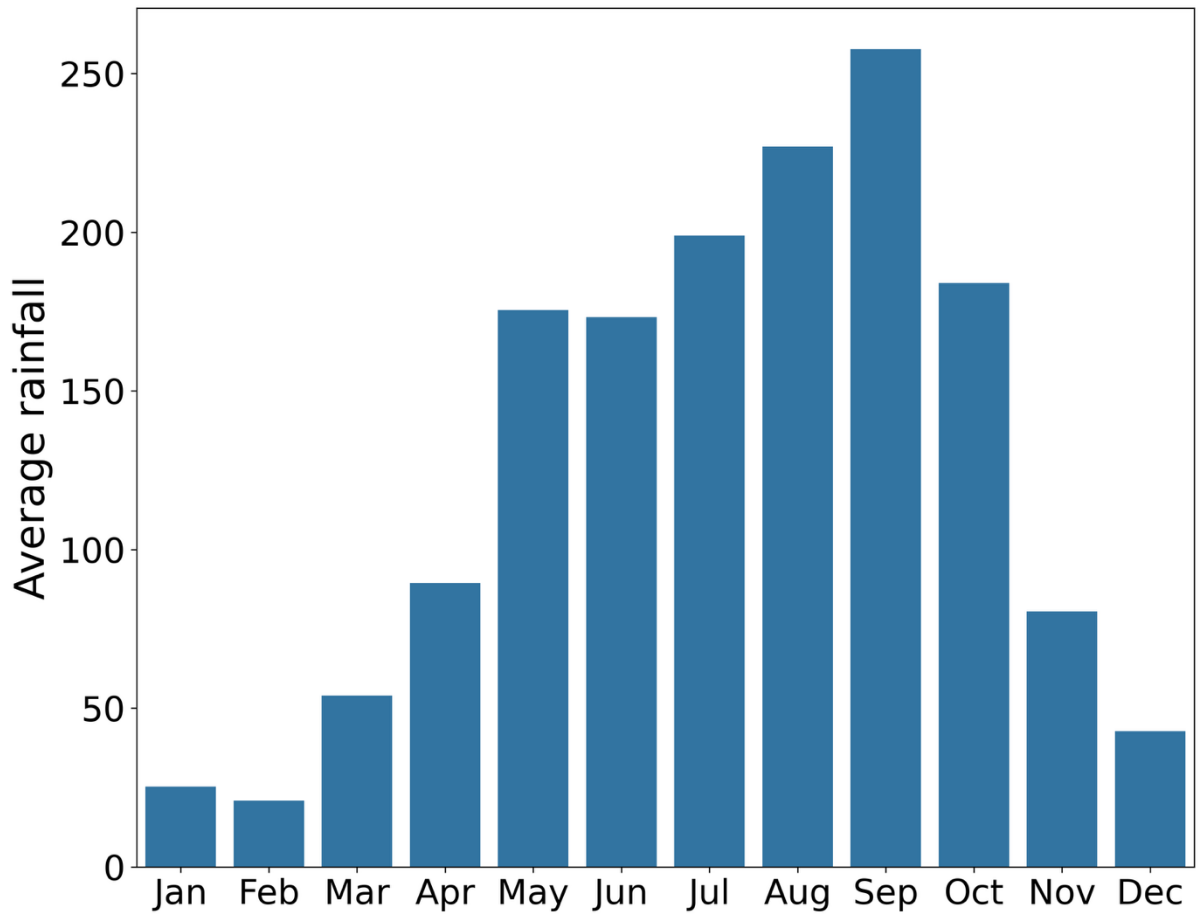


**Figure S5. Monthly average rainfall patterns across Thailand, 2003-2022.** Bars represent mean monthly precipitation (mm) averaged across all 77 provinces over the 20-year study period. Data derived from the TerraClimate dataset. Note the peak rainfall in August-September contrasts with peak dengue transmission in July (see Figure S1), supporting a role of precipitation in a complicated non-monotonic fashion in dengue risk prediction.
